# What does 16S rRNA gene-targeted next generation sequencing contribute to the study of infective endocarditis in valve tissue?

**DOI:** 10.1101/2021.01.23.21250364

**Authors:** Paula Santibáñez, Aránzazu Portillo, Sonia Santibáñez, Lara García-Álvarez, María de Toro, José A. Oteo

**Author notes:** Address correspondence to: José A. Oteo, Hospital Universitario San Pedro-CIBIR. Infectious Diseases Department. Center of Rickettsiosis and Arthropod-borne diseases., C/ Piqueras, 98 26006 – Logroño (La Rioja) SPAIN.

## Abstract

Infective endocarditis (IE) is a severe and life-threatening disease. Identification of infectious etiology is essential for establishing the appropriate antimicrobial treatment and decreasing mortality. The aim of this study was to explore potential utility of metagenomics for improving microbiological diagnosis of IE. In this work, next-generation sequencing (NGS) of V3-V4 region of the 16S rRNA gene was performed in 27 heart-valve tissues (18 natives, 5 intravascular devices, and 4 prosthetics) of patients diagnosed by IE. Initial microbiological diagnosis, blood culture (BC) and/or PCR, was compared with NGS-based diagnosis. Metagenomics matched with conventional techniques diagnosis in 24/27 cases (88.9%). The same bacterial family was assigned to 24 cases, the same genus to 23 cases, and the same specie for 13 cases. In 22 of them, the etiological agent was represented by percentages >99% of the reads and in two by ∼70%. Staphylococcus aureus was detected in a previously undiagnosed patient, making the microbiological diagnosis possible in one more sample than with previously used techniques. The remaining two patients showed no coincidence between traditional and NGS microbiological diagnoses. Minority records verified mixed infections in four cases and suggested confections in two cases, supported by clinical data. In conclusion: 16S rRNA gene-targeted NGS allowed to diagnose one case of IE without microbiological entity based on traditional techniques. However, the application of metagenomics to the study of IE in resected heart valves provides no benefits in comparison with BC and/or PCR. More studies are needed before implementation of NGS for the diagnosis of IE.

## Text

### Introduction

Infective endocarditis (IE) is defined as an infection of a native or prosthetic cardiac valve, endocardial surface, or indwelling cardiac device (1).Despite trends towards earlier diagnosis, pharmacotherapy, and surgical intervention, IE remains a major medical concern with an unchanged incidence (1.7-10 cases/100,000 inhabitants) and mortality (13%-25% in-hospital, approaching 40% within the first year) in the last two decades (2, 3).

Identification of the causative agent/s is essential for optimal patient management and guides treatment duration and antibiotic choice.Microbiological diagnosis mainly relies on cultured-based methods (4, 5).However, blood cultures are negative in 2.5%-31% of IE cases (3, 6).Causes of blood culture negative endocarditis (BCNE) include sub-optimal specimen collection, antibiotic treatment prior to blood culture collection, and/or infection due to fastidious microorganisms or intra-cellular bacteria (e.g.deficient streptococci, members of the HACEK group, *Propionibacterium acnes, Candida* sp., *Brucella* spp., *Legionella* spp., *Listeria monocytogenes, Mycoplasma* spp., mycobacteria, *Coxiella burnetii, Bartonella* spp., *Tropheryma whipplei*, etc.) (2, 7).These limitations are overcome by molecular biology techniques.Thus, specific and broad range 16S rRNA PCR assays from blood and/or valvular biopsies have proven to be useful for etiologic diagnosis (6, 8, 9).However, heart valves are not usually available, and they are only after surgery, which may delay an early diagnosis.

In the last two decades, next-generation sequencing (NGS) technology has been extensively used in research studies, particularly focusing on the human microbiota and its association with health and disease (10).Continuous improvements of NGS technology as, well as the reduction of costs, have effectively transformed biomedical research (11-13).Nowadays, clinical laboratories evaluate metagenomics as an infectious diseases diagnostic tool for identification of microorganisms directly from patient samples (14).

The aim of this work was to evaluate the contribution of targeted 16S rRNA gene-NGS analysis in heart valve tissues to the study of IE.

## Materials and Methods

### Samples

DNA from resected heart valves extracted using QIAamp DNA Mini Kit (Qiagen, Hilden, Germany) and stored at -80ºC, were retrospectively selected from the CRETAV collection (CIBIR, La Rioja, Spain).They corresponded to 18 native heart valves, five intravascular devices, and four prosthetic heart valves from 27 patients diagnosed with IE according to the modified Duke criteria in our hospital (Hospital Universitario San Pedro, La Rioja, Spain) from 2009 to 2017 (Table 1).Clinical data and results of blood culture (BC) and 16S ribosomal RNA (rRNA) gene PCR (15) and Sanger sequencing from the heart valve tissues were available in all cases (Table 1).

**Table 1:**
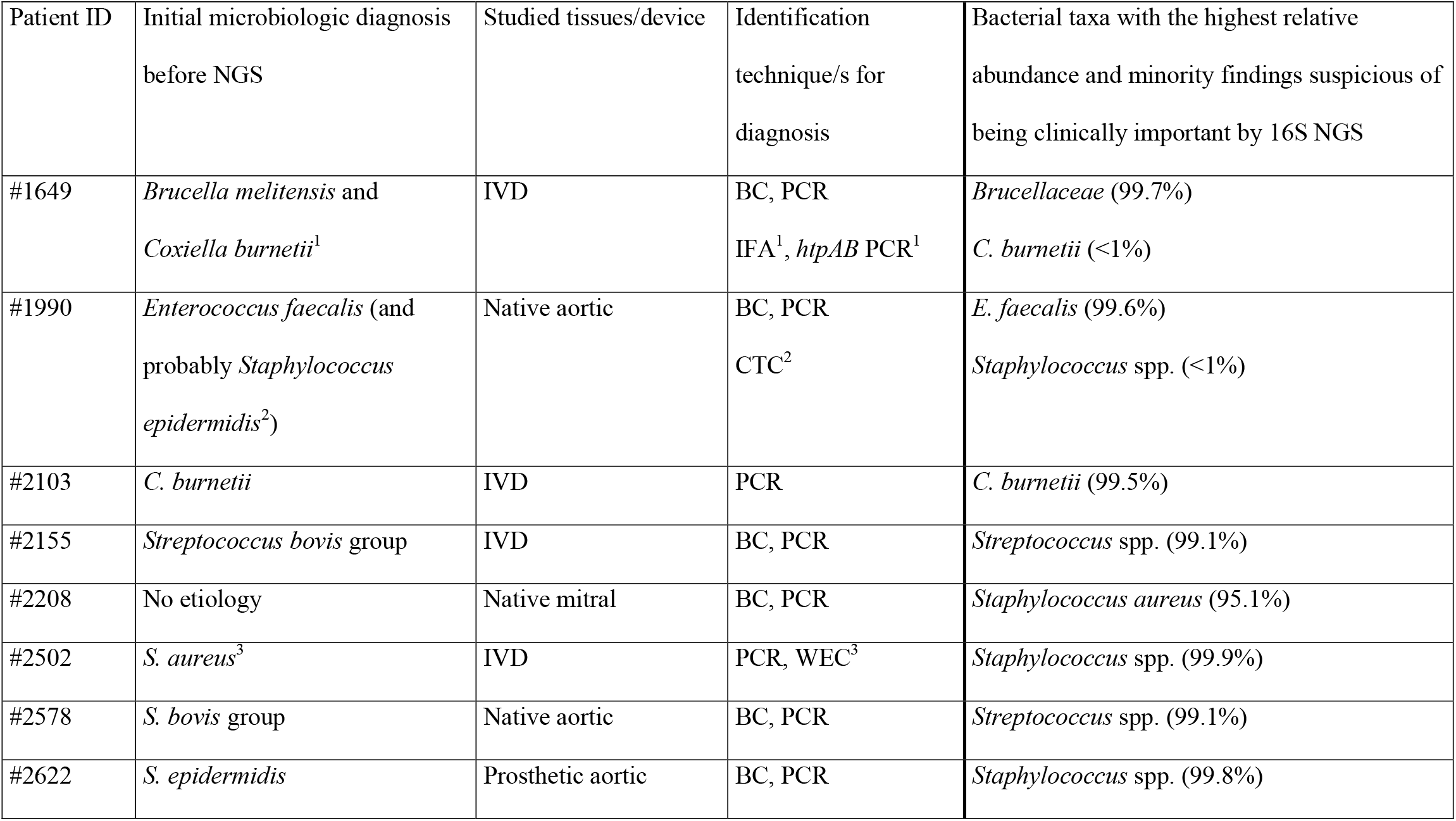
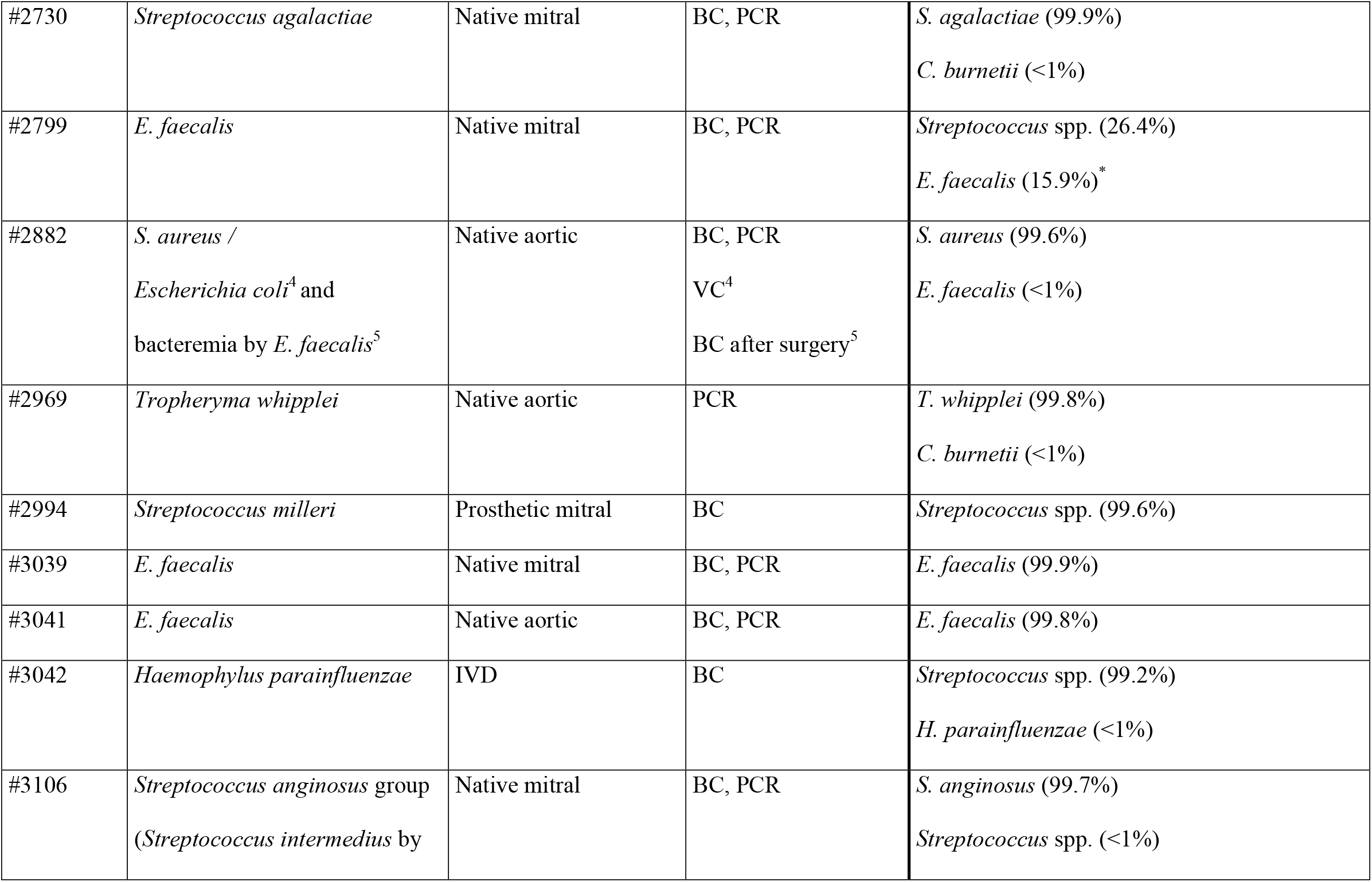
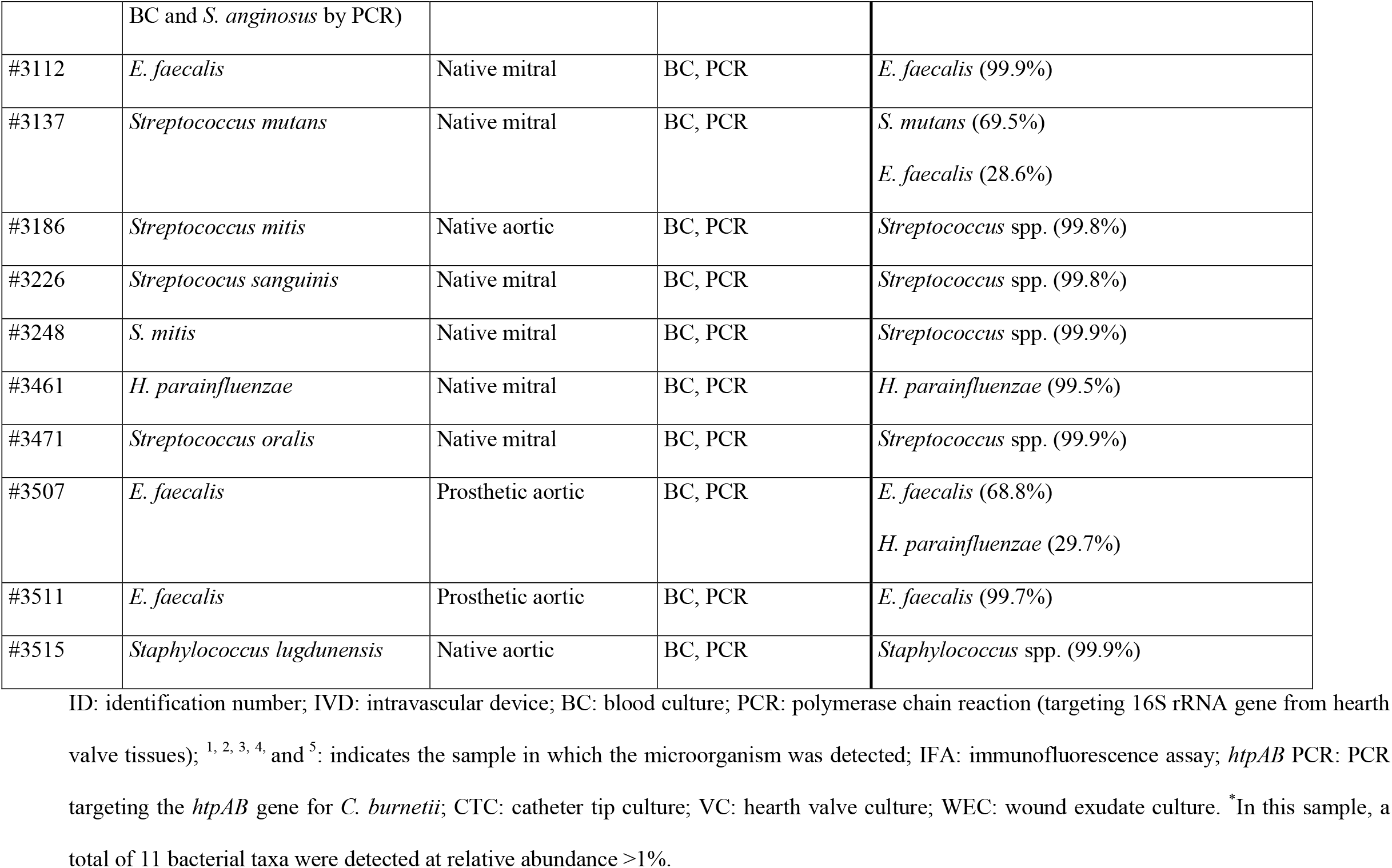
Contributions of NGS analysis from hearth valve tissue specimens to the diagnosis of infective endocarditis (IE) for the 27 patients of this study.

Approval of the regional ethics committee was obtained (Comité Ético de Investigación Clínica-Consejería de Sanidad de La Rioja, Ref.CEICLAR PI-19).Informed consent was obtained from all participants.All procedures were in accordance with the ethical standards of the research committee and with the 1964 Helsinki declaration and its later amendments.

### DNA quantification and quality determination

DNA was quantified with a Qubit 3.0 fluorometer (Thermo Scientific) using Qubit dsDNA HS (High Sensitivity) assay kit.The quality of DNA was assessed with the Fragment Analyzer by Agilent formerly Advanced Analytical (AATI) using Genomic DNA 50kb kit.A total of 12.5 ng DNA per sample were added.

Samples were manipulated under sterile conditions in a Class II biosafety cabinet using cycles of UV light prior and between uses to prevent contamination.Sterile single-use instruments were used.DNA extraction, preparation of PCR master mix, and amplification were performed in separate rooms to prevent contamination.

### 16S rRNA gene amplification, library preparation and sequencing

Primers targeting the hypervariable V3 and V4 regions of 16S rRNA gene were used (16).Amplified regions were purified and indexed with Nextera XT Index kit (Illumina).The library quality was assessed on a Qubit 3.0 Fluorometer (Thermo Scientific) and Fragment Analyzer (AATI) using dsDNA reagent (35-5000bp) kit.Paired-end 300 bp sequences were obtained on an Illumina MiSeq platform.

### Sequence processing and analysis

Quality controls of raw reads were carried out with FastQC software http://www.bioinformatics.babraham.ac.uk/projects/fastqc (17), and trimmed with the Trimmomatic software (18).The V3-V4 amplified region (550-580 bp) was reconstructed through paired reads according the Quantitative Insights Into Microbial Ecology (QIIME) protocol (v1.9.1) (19).Operational Taxonomic Units (OTUs) were defined as sequences with at least 97% similarity versus Greengenes database (20) using UClust clustering algorithm http://drive5.com/usearch/manual/uclust_algo.html (21) and following the open-reference method described by QIIME (22).OTUs with <0.01% relative abundance of the total read counts on a per-sample basis were removed (spurious and chimeric reads).Each OTU’s sequences were refined with BLAST tool against GenBank https://blast.ncbi.nlm.nih.gov/Blast.cgi?PROGRAM=blastn&PAGE_TYPE=BlastSearch&LINK_LOC=blasthome# (23).The causative pathogen was attributed to the microorganism with which the most amplicon reads matched.Low-abundant taxa were also considered relevant when a diagnosis with mixed infections matched epidemiologic, clinical and microbiologic results of patients.

## Results

The 16S rDNA gene amplicon sequencing successfully revealed bacterial pathogens in all samples of the cohort (Table 1).A total of 8,733,978 reads [average counts per sample 323,481; minimum: 67,043 and maximum: 672,563] distributed among 118 OTUs were obtained.The metagenomic analysis of bacteria in hearth valve tissues revealed the same microbiologic diagnosis than conventional techniques (BC and/or PCR) in 24 out of 27 cases.However, the accuracy of the 16S-NGS test did not enable us to give all these results at the level of species.With this technology, it was possible to assign the same bacterial family as with traditional techniques for 24 cases, the same genus for 23 cases, and the same species for 13 cases.For 22 out of these 24 cases that showed consistent results among NGS data and conventional methods, the bacterial taxa with maximum representation were found in a proportion of reads that ranged 99.1-99.9%.However, for two cases (patient IDs #3137 and #3507), around 70% of the reads confirmed previous results (*S*.*mutans* and *E*.*faecalis*, respectively) whereas reads at nearly 30% corresponded to *E*.*faecalis* and *H*.*parainfluenzae*, respectively.For all these 24 patients, NGS results allowed us to retrospectively confirm the previous diagnosis.In addition, the 16S-NGS test data assessment, led us to corroborate mixed infections within the clinical and epidemiologic context when considering relatively low abundant microorganisms.Thus, for patient ID #1649, whose initial microbiologic diagnosis included *Brucella melitensis* (by BC and PCR) and *Coxiella burnetii* (by indirect immunofluorescence assays and specific PCR) (24), metagenomic results showed *Brucellaceae* as the most relative abundant taxa (99.7%) and, interestingly, *C*.*burnetii* was also detected at low relative abundance (<1%).For patient ID #1990, conventional methods (BC and PCR) led to an IE diagnosis due to *E*.*faecalis*, and probably to *S*.*epidermidis* since this bacterium was obtained from the culture of the tip of the catheter.According to the 16S rRNA gene profiling analysis, 99.6% of reads mapped to *E*.*faecalis*, and members of the genus *Staphylococcus* were also observed (<1%).For patient ID #2882, an initial diagnosis with IE by *Staphylococcus aureus* was given according to BC and PCR methods.Furthermore, culture of the resected hearth valve tissue yielded *Escherichia coli*, and bacteremia by *E*.*faecalis* was detected by BC after surgery.The 16S-NGS work revealed that the majority of the reads (99.6%) corresponded to *S*.*aureus*, and *E*.*faecalis* was also present in a lower proportion of reads (<1%).In this case, no sequenced reads mapping to *E*.*coli* were obtained.Patient ID #3106 was diagnosed with IE by viridans group streptococci based on traditional techniques (BC yielded *Streptococcus intermedius* and *Streptococcus anginosus* was detected by PCR).According to the metagenomic analysis of this specimen, the percentage of sequenced reads observed matching *S*.*anginosus* reached 99.7%, and other streptococci including *Streptococcus sanguinis, Streptococcus cristatus* and *Streptococcus mutans* were found with low relative abundance (<1%).

In addition, two cases were reclassified as mixed infections since sequence reads of uncultured microorganisms were detected and they were subsequently assessed in the clinical-epidemiologic context.NGS test results from patient ID #2730 showed high relative abundance of *S*.*agalactiae* (99.9%) and low relative abundance of *Coxiellaceae* (<1%).These data corresponded to a patient diagnosed with IE by *S*.*agalactiae* by BC and PCR, with *C*.*burnetii* phase II IgG titer of 400 and phase I IgG not detected, with negative PCR results for *C*.*burnetii*, and positive PCRs for bacteria within the *Coxiellaceae* family.For the other case (patient ID #2969), diagnosed with IE by *T*.*whipplei* based on PCR results, the NGS analysis showed *T*.*whipplei* as the most abundant bacteria (99.8%) and <1 % of reads were assigned to *Coxiellaceae*, suggesting a mixed infection supported by serological criteria (*C*.*burnetii* phase I IgG titer ≥1,600 and phase II IgG titer of 800).

For the sample in which no microbiologic diagnosis had been achieved either by BC or by PCR (patient ID #2208), the analysis of metagenomics sequences evidenced that most of them (95.1% of relative abundance) corresponded to *S*.*aureus*.Thus, with our NGS approach we were able to detect the causative agent of IE in one more sample than with the remaining methods.

In contrast, the testing by V3-V4 16S metagenomics yielded taxonomic predictions that differed from those obtained with conventional methods for two cases.Nevertheless, according to our NGS data, the clinical entities found with conventional techniques were also detected in these two samples, although at low relative amounts (patient IDs 182 #2799 and #3042).

Metagenomic results corresponding to the remaining 16 patients not detailed above were consistent with those obtained by traditional techniques, and NGS technology did not provide additional information considered relevant for diagnosis.

Detailed information about data that support the diagnosis of the studied patients is showed in Table 1.

## Discussion

IE is still a severe disease with high morbidity and prolonged hospital stay as well as very high mortality during admission and during the 1-year follow-up (3).Therefore, techniques to reliably guide the correct antimicrobial treatment in order to get the sterilization of the affected tissues and decrease the mortality are needed (8).NGS has recently emerged as a comprehensive method for exploring causative agents of infectious diseases without prior culture.However, reports about metagenomic analysis for pathogen identification in hearth valve tissues are scarce, and each of them consists of very few patients (25-34).Here, we used 16S rRNA targeted-NGS from hearth valve tissues as an approach to the diagnosis of IE in a cohort of 27 patients.

Herein, metagenomics allowed the microbiological diagnosis (*S*.*aureus*) in patient #2208, in which the causative agent had not been detected either by PCR or by HC.In addition, the same microbiological diagnosis was obtained using NGS or routine techniques for 24 patients (88.9%), although at higher taxonomical level for 11 of them.Taxonomic assignment of sequence reads below the genus level is a challenge in metagenomics data analysis.Only the combination of multiple hypervariable regions or the nearly complete sequence of 16S rRNA gene gives accurate measures of taxonomic diversity (35).Third-generation sequencing provides long-read sequences, but high base-calling error rates (36).Besides, consensus in current NGS protocols is essential since microbiome studies are potentially biased at every methodological stages, from sampling to bioinformatic analysis (37, 38).

The interpretation of the metagenomic results for IE cases has been based on considering the bacteria represented by the highest percentage of reads as the causative agent (26, 30, 32).In our series, this was possible for 22 patients with values from 99.1% to 99.9% of the reads and in 1 patient (patient ID #2208) with 95.1% of the reads.However, we found two cases (patient IDs #3137 and #3507) in which the most represented bacteria matched to the initial causative agent, but the proportion reached 70% of the reads, and the remaining ones corresponded to *E*.*faecalis* and *H*.*parainfluenzae*, respectively.The biological significance of reads at around 30% of relative abundance is currently unknown.Moreover, in patient ID #2799 the highest proportion of reads was as low as 26.4% and mapped to *Streptococcus* spp., and other 10 bacterial taxa were detected at relative abundance >1% (Supplementary data).

One main advantage of 16S rRNA-NGS technology is its capacity to classify all bacteria from a sample without intermediate culturing steps (14).Minority findings constitute a concern for 16S rRNA gene NGS projects.The cutoff value indicating how many reads of a microorganism in a sample are no relevant for the analysis has not been established for microbiome studies.It will require a vast experience to establish which are spurious reagent contaminants, sample processing contaminants, cross-contamination in multiplexed libraries, etc.or true infections or coinfections.Regarding to the relatively low abundant microorganisms, we have described four cases of previously characterized mixed infections (patient IDs #1649, #1990, #2882 and #3106) and two cases reclassified as mixed infections without clinical suspicion, but in agreement with serological results (patient IDs #2969 and #2730).However, there were two cases in which the microorganism formerly considered as causative agent by HC and/or PCR was barely represented in metagenomic results (patients IDs #2799 and #3042).In order to understand this, it is important to take into consideration that, as was previously mentioned, each step of NGS analysis influences the relative abundances observed (36-38).However, it cannot be discarded that these small percentages are inherent failures of the technique.The concordance of minority percentages with clinical data gave value to the diagnosis of mixed infectious in the six cases mentioned above (patient IDs #1649, #1990, #2882, #3106 #2969 and #2730), in contrast to patient ID #3042, in which no clinical or epidemiological data available supported streptococcal infection, and patient ID #2799, in which 11 bacterial taxa were found.Addressing laboratory contamination is an urgent task and it is important to scrutinize NGS data with an understanding of its potential for false positive results.Bacterial identification using 16S rRNA-NGS may be biased because of unequal amplification of certain species, and it is influenced by several factors, such as the region(s) sequenced, amplification efficiency, sequencing technology, and bioinformatics workflow(s).Including spiking-in mock microorganisms that provide comparable results across research groups and time, as well as positive and negative controls in order to assure the quality of the NGS results, is recommended (36, 38).

NGS technology has been suggested as an essential supplement to culture-based methods for the diagnosis of IE, particularly for unculturable or difficult-to-culture microorganisms (25, 26 31, 32).However, in our three BCNE cases, the microbiological diagnosis had already been achieved by 16S rRNA PCR (patient IDs #2103, #2502 and #2969).Therefore, the application of NGS techniques may not always be valuable, even more considering that is more expensive and time-consuming, and requires equipment hardly affordable by most clinical laboratories and personnel trained in bioinformatics analysis.

Anyway, this is so far one of the largest series published, and the concordance of our results with the previous microbiological diagnoses in almost all patients highlights the importance of this work.

## Conclusions

Results of 16S rRNA-NGS are mostly consistent with those of BC and/or conventional PCR but do not improve the diagnosis of IE cases.Metagenomics may be helpful to IE patients after valve replacement surgery, especially when conventional tests fail to yield a diagnosis.Moreover, minority findings supported by clinical data could suggest mixed infections not previously suspected, although more efforts should be made in order to understand them.Hence, further studies are required to validate the clinical usefulness of this method.

## Data Availability

All data referred to in the manuscript are available.

## Acknowledgments

To María Bea, laboratory technician from the Genomics and Bioinformatics Core Facility (CIBIR, La Rioja, Spain).

Partial results were presented at VII Congreso SEICAV (Sociedad Española de Infecciones Cardiovasculares), Sevilla, 2018.

## Transparency declaration

The authors have no conflicts of interest to disclose.No specific external funding was available for this study.

## Supplementary data

| Bacteria | IE agent | Clinical importance | Reference |
| --- | --- | --- | --- |
| *Abiotrophia defectiva* | <1% | Normal flora of the oral cavity, the urogenital and the intestinal tracts. It has been involved in ocular, osteoarticular, arthroplasty and sinus infections, otitis, cerebral abscesses, iatrogenic meningitis and pancreatic abscesses. | Birlutiu et al., 2017 |
| *Actinomyces* spp. | 30 cases since 1939 | Infections are usually limited to cervicofacial, thoracic, and abdominopelvic regions. | Phichaphop et al., 2020 |
| *Bergeyella zoohelcum* | rare | It is a well-known zoonotic pathogen causing cellulitis, leg abscess, tenosynovitis, septicemia, pneumonia and meningitis. | Pan et al., 2020 |
| *Capnocytophaga sputigena* | yes | Normal flora of the oral cavity. Has occasionally been associated with periodontitis, sinusitis, conjunctivitis, pleural empyemas, subphrenic abscesses, osteomyelitis and septicemia. | Torru et al., 1995 |
| *Corynebacterium* spp. | yes | Normal skin flora. Contaminate blood cultures and cause opportunistic severe infections. | Rasmussen et al., 2020 |
| *Enterococcus faecalis* | 10%<br>(Enterococci) | Part of the human gastrointestinal flora. Cause urinary, intra-abdominal, wound and pelvic infections and bacteremia. It is the third most common cause of IE. | Dahl et al., 2013 |
| *Haemophilus parainfluenzae* | 1.2 - 3% (HACEK group) | Frequently colonize the oropharynx and nasopharynx and may also be part of the genital tract microbiota. Related to infections of the respiratory tract although brain abscess, surgical site infections, soft tissue infections, prosthetic joint infections, and hepatic and biliary tract infections have also been documented. Most common cause of | Revest et al., 2016 |
|  |  | HACEK endocarditis. |  |
| *Lautropia mirabilis* | no | Commonly found in the human oral cavity and the upper respiratory tract. Described in HIV-positive children and in the sputum of one cystic fibrosis patient. | Rossmann et al., 1998;<br>Voronina et al., 2018 |
| *Neisseria elongata* | very rare | Normal oropharyngeal bacterial flora. Associated with septicemia and osteomyelitis. | Youssef et al., 2019 |
| *Rothia aeria* | 9 cases | Colonizes the human oral cavity and upper gastrointestinal tract. Sporadically related as cause of neck abscess, shoulder joint infection, lung infection and neonatal sepsis. | Kim et al., 2014 |
| *Streptococcus* spp. | 30% | Streptococci are found in almost every location in the human body and are the dominant species in the human oral cavity and upper respiratory tract. They are the second leading cause of IE. | Kim et al., 2018 |

## Notes

### Competing Interest Statement

The authors have declared no competing interest.

### Author Declarations

Approval of the regional ethics committee was obtained (Comite Etico de Investigacion Clinica-Consejeria de Sanidad de La Rioja, Ref. CEICLAR PI-19). Informed consent was obtained from all participants. All procedures were in accordance with the ethical standards of the research committee and with the 1964 Helsinki declaration and its later amendments.

